# Development of population and Bayesian models for applied use in patients receiving cefepime

**DOI:** 10.1101/19009647

**Authors:** Jiajun Liu, Michael Neely, Jeffrey Lipman, Fekade Sime, Jason Roberts, Patrick J Kiel, Sean N. Avedissian, Nathaniel J. Rhodes, Marc H. Scheetz

**Affiliations:** Midwestern University Chicago College of Pharmacy, Downers Grove, IL USA; Northwestern Memorial Hospital, Chicago, IL USA; Midwestern University Chicago College of Pharmacy Pharmacometrics Center of Excellence, Downers Grove, IL USA; Children’s Hospital Los Angeles and University of Southern California, Los Angeles, CA; UQ Centre for Clinical Research, Faculty of Medicine, University of Queensland, Australia; Royal Brisbane and Women’s Hospital, Brisbane, Australia; Centre for Translational Anti-infective Pharmacodynamics, School of Pharmacy, University of Queensland, Australia; Indiana University Health, Indianapolis, IN, USA; Antiviral Pharmacology Laboratory, University of Nebraska Medical Center (UNMC) Center for Drug Discovery, UNMC, Omaha, NE USA; University of Nebraska Medical Center, College of Pharmacy, Omaha, NE USA

**Keywords:** cefepime, pharmacokinetic, pharmacodynamic, individualized dosing, precision dosing, model, population model

## Abstract

Understanding exposures of cefepime, a β-lactam antibiotic, is crucial for developing regimens to achieve optimal exposure and improved clinical outcomes. This study sought to develop and evaluate a unified population pharmacokinetic model in both pediatric and adult patients receiving cefepime treatment. Multiple physiologically relevant models were fit to pediatric and adult subject data. To evaluate the final model performance, a withheld group of twelve pediatric and two separate adult populations were assessed. Seventy subjects with a total of 604 cefepime concentrations were included in this study. All adults (n=34) on average weighed 82.7 kg and displayed a mean creatinine clearance (CrCL) of 106.7 mL/min. All pediatric subjects (n=36) had mean weight and CrCL of 16.0 kg and 195.64 mL/min, respectively. A covariate-adjusted two compartment model described the observed concentrations well (population model R^2^, 87.0%; Bayesian model R^2^, 96.5%). In the evaluation subsets, the model performed similarly well (population R^2^, 84.0%; Bayesian R^2^, 90.2%). The identified model serves well for population dosing and as a Bayesian prior for precision dosing.

**Key points:**

- A unified cefepime population pharmacokinetic model has been developed from adult and pediatric patients and evaluates well in independent populations.
- When paired with real time beta-lactam assays, precision dosing approach will optimize drug exposure and improve clinical outcomes.

## 1. Introduction

Cefepime is a commonly utilized antibiotic for nosocomial infections. Rising resistance, manifesting as increased cefepime minimum inhibition concentrations (MICs), has led to more frequent clinical failures [1, 2]. In order to advise clinical outcomes according to MIC, the Clinical and Laboratory Standards Institute (CLSI) updated the susceptibility breakpoints and then created a category of susceptible-dose dependent for MICs of 4 and 8 mg/L for *Enterobacteriaceae spp*. [3]. Achieving goal pharmacokinetic exposures to effectively treat these higher MICs can require a precision dosing approach.

Cefepime, like other β-lactams, has pharmacodynamic activity governed by ‘time-dependent’ activity. The fraction of time that the unbound drug concentration exceeds the MIC (*f*T_>MIC_) for the dosing interval is the pharmacokinetic/pharmacodynamic (PK/PD) efficacy target for cefepime [4], and a target of 68%-74% has been established [5]. For the currently approved cefepime product and combination agents in the pipeline [6, 7], understanding cefepime disposition and variability is crucial to optimally treat the patients. Since inter- and intra-patient pharmacokinetic variability can impact the achievement of pharmacodynamic goals, understanding the precision of population dosing is important. Further, to fully realize precision dosing, individualized models (e.g. Bayesian models) are needed. Once developed, these models will form the basis for adaptive feedback and control strategies when paired with real time drug assays.

The purpose of this study was to: 1) develop and evaluate a unified cefepime population PK model for adult and pediatric patients, and 2) construct an individualized model that can be utilized to deliver precision cefepime dosing.

## 2. Material and methods

### 2.1 Study Populations

Data from four clinical cefepime PK studies representing unique groups of patients were compiled. Subject demographics and study methodologies have been previously described [8-11]. In brief, populations represented were febrile neutropenic adults with hematologic malignancies [10, 11], those with critical illness [9] and children with presumed or documented bacterial infections [8]. For the two studies that evaluated adults with neutropenic fever, Sime et al. prospectively enrolled 12 patients receiving chemotherapy and/or stem cell transplant who subsequently developed febrile neutropenia and were administered maximum doses of cefepime [10]. A total of 53 cefepime plasma concentrations in presumably steady-state dosing intervals (third, sixth, and ninth) were analyzed for PK target attainment. Whited et al. prospectively studied similar patients (n=9) who were admitted to hematology-oncology services and were receiving cefepime at maximum dosage for febrile neutropenia [11]. Cefepime PK samples were obtained during steady state and analyzed for population parameters. Critically ill adults were studied by Roberts et al. as a prospective multinational PK study and included 14 patients who received cefepime [9]. Lastly, Reed et al. characterized cefepime PK in hospitalized pediatric patients (above 2 months of age) who received cefepime as monotherapy for bacterial infections [8]. For our study, only those who received intravenous cefepime were included for model development.

### 2.2 Model Building Populations

Adult (n=34) and partial pediatric (n=36) subjects were utilized for PK model building (Fig. 1) while model evaluation was performed with other datasets consisting of independent adult (n=25) and pediatric (n=12) patients. Pediatric patients from Reed et al. [8] were randomized into the model building or the evaluation dataset. All clinical patient level data included age, weight, and serum creatinine (SCr). An estimated creatinine clearance (CrCL) was calculated for each patient [12]. The Cockcroft Gault formula served as a standardized descriptor for elimination rate constant (Supplemental Fig. 1). This study was exempted by the Institutional Review Board at Midwestern University Chicago College of Pharmacy.

### 2.3 Pharmacokinetic Models

To construct the base PK models, the Nonparametric Adaptive Grid (NPAG) algorithm [13, 14] within the Pmetrics (Version 1.5.2) package [14] for R [15] was utilized. Multiple physiologically relevant one- and two compartmental PK models were built and assessed. The one-compartment structural model included an intravenous cefepime dose into and parameterized total cefepime elimination (K_e_) from the central compartment (V_C_). The two-compartment model included additional parameterizations of intercompartmental transfer constants between central and peripheral compartments (K_CP and_ K_PC_). In candidate models, total cefepime elimination was explored according to full renal and partial renal clearance (CL) models [i.e. non-renal elimination (K_e_Intcpt) and renal elimination descriptor (K_e_0 vectorized as a function of glomerular filtration estimates)] [6, 16].

**Figure 1.**
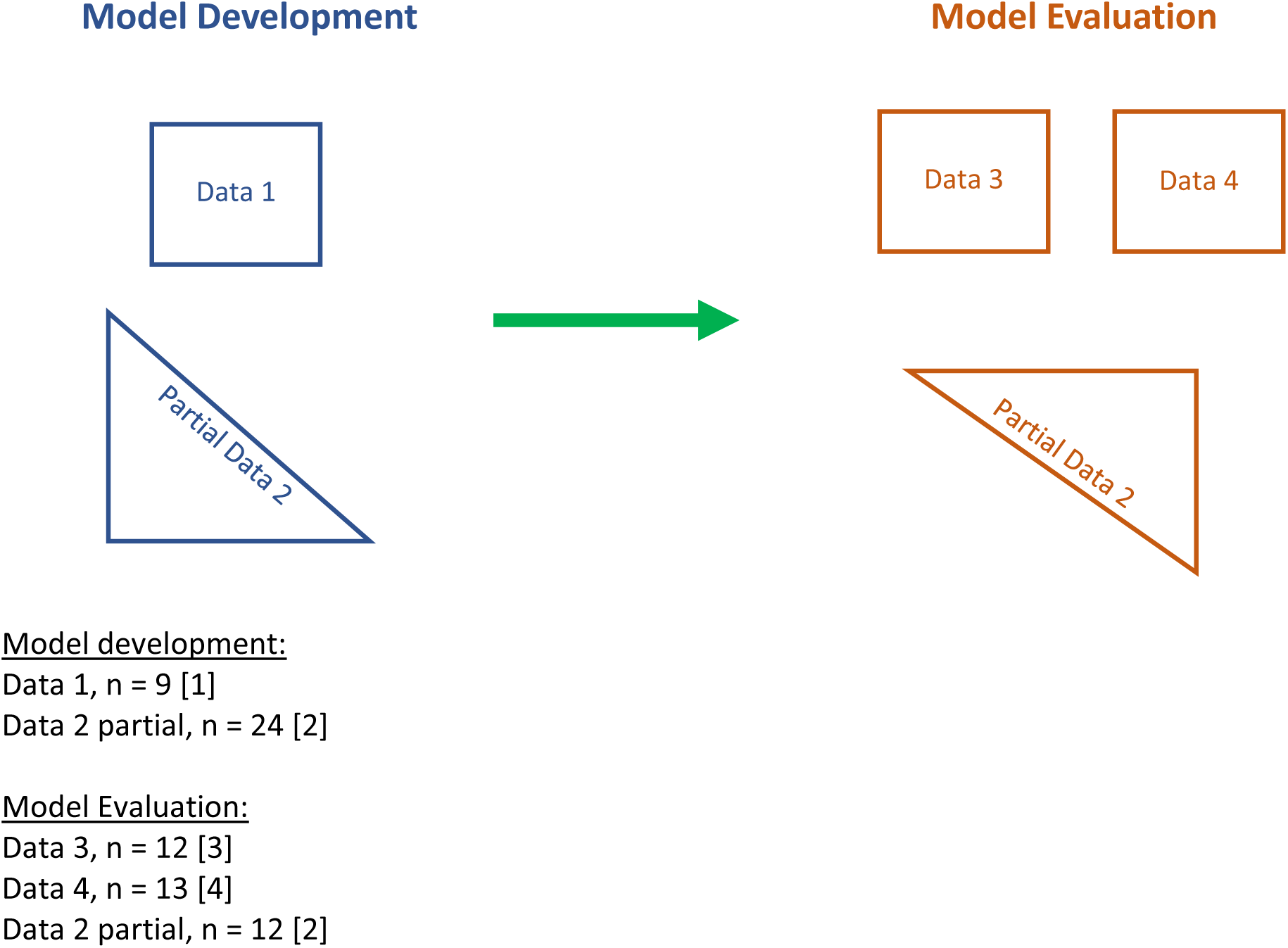
Schematic for Data Sources in Model Development and Evaluation.

Assay error was included into the model using a polynomial equation in the form of standard deviation (SD) as a function of each observed concentration, Y (i.e. SD = C_0_ + C_1_ · Y). Observation weighting was performed using gamma (i.e. error = SD · gamma), a multiplicative variance model to account for extra process noise. Gamma was initially set at 4 with C_0_ and C_1_ equal to 0.5 and 0.15, respectively.

Covariate relationships were assessed using the ‘PMStep’ function in Pmetrics by applying stepwise linear regressions (forward selection and backwards elimination) of all covariates on PK parameters. Additionally, *a priori* analyses examined the effect of covariates (e.g. weight, CrCL) on cefepime elimination rate constant (K_e_) because of their known importance in describing cefepime disposition [6, 17, 18]. Weight and CrCL were standardized to 70 kilogram (kg) and 120 mL/min, respectively. Further, an allometric scaler was applied to standardized weight (i.e. (quotient of weight in kg divided by 70 kg raised to the 0.75^th^ power) as a covariate adjustment to K_e_ (Supplemental Fig. 1). Ultimate model retention was governed according to criteria described below.

The best structural and error model was identified by the change in objective function value (OFV) calculated as differences in −2 log-likelihood (−2LL), with a reduction of 3.84 in OFV corresponding to p <0.05 based on chi-square distribution and one degree of freedom. Further, the best-fit model was selected based on rule of parsimony and the lowest Akaike’s information criterion (AIC) scores. Goodness-of-fit of the competing models were evaluated by regression on observed vs. predicted plots, coefficients of determination, and visual predictive checks. Predictive performance was assessed using bias and imprecision in both population and individual prediction models. Bias was defined as mean weighted prediction error; imprecision was defined as bias-adjusted mean weighted squared prediction error. Posterior-predicted cefepime concentrations for each study subject was calculated using individual median Bayesian posterior parameter estimates. Using Bayesian posterior-predicted concentrations (at every 0.2 h) from the final model, noncompartmental analyses (NCA) were performed to estimate additional PK parameters [i.e. half-life, clearance (CL)].

### 2.4 Model Evaluation

To evaluate the final adjusted model, the NPAG algorithm [13, 14] was employed to assess the performance with separate data sets (Fig. 1). The population joint density from the final adjusted model were employed as Bayesian prior for the randomly withheld pediatric and adult data. In the evaluation process, structural model, model parameters, assay error, and observation weighting were unchanged. Goodness-of-fit of the competing models were determined as described above.

### 2.5 Simulations and probability of target attainment (PTA)

Simulation was performed to examine the exposures predicted by the final adjusted PK model [14, 19]. Covariate values were fixed per subject based on arithmetic means of observed weight and CrCL for adult and pediatric in the model development populations. Monte Carlo sampling was performed from the multimodal, multivariate distribution of parameters with limits fixed by the bounded parameter space. Maximum dosing regimens were simulated for adult and pediatric populations: 2 grams every 8 hours (h) infused over 0.5 h and 50 mg/kg every 8 h infused over 0.5 h. Protein binding of 20% (i.e. 80% free fraction of total cefepime dose) was accounted for in predicting 48-h cefepime concentrations [6]. For each scenario, 2,000 parameter sets or patients were generated. PK/PD target of *f*T_>MIC_ ≥ 68% was utilized across doubling MICs of 0.25-32 mg/L over the first 48 h of cefepime therapy [5]. Estimates are provided from the first 24 h of simulations as timely administration of effective antimicrobial agents is associated with increased survival [20].

## 3. Results

### 3.1 Demographics

A total of 70 clinically diverse subjects, contributing 683 cefepime concentrations, were included in this study (n=45 subjects for model development; n=25 subjects for evaluation). Adult subjects (n=34) had a mean weight [SD] of 82.7 kg [21.5] and mean CrCL [SD] of 106.7 mL/min [58.4]. For the pediatric cohort (n=36), means [SD] of weight and CrCL were 16.0 kg [16.1] and 195.64 mL/min [40.5], respectively. Adult subjects ranged in age from 22-82 years (mean, 55.4 years) while pediatric subjects ranged from approximately 2 months to 16 years of age (mean, 3.9 years).

### 3.2 PK model selection, parameters, and evaluation

A total of 604 cefepime observations were available for analysis. Cefepime concentrations ranged from 0.5-249.7 μg/mL. The base one- and two-compartment models (no covariate adjustment) produced reasonable fits for observed and Bayesian posterior-predicted cefepime concentrations (R^2^=84.7% and 85.2%, respectively), but population estimates were unsatisfactory (R^2^=22.7% and 27.8%, respectively) (Table 1). After standardizing weight (to 70 kg) without an allometric scaler in the base two-compartment model, fits for both population and Bayesian posterior estimates against the observed data improved (R^2^=60.7% and 96.5%, respectively; OFV change, 4). Bias and imprecision for Bayesian posterior fits were −0.18 and 1.12, respectively. When covariates (i.e. weight to volume of distribution and K_e_; CrCL to K_e_) and the allometric scaler were applied in the two-compartment model, Bayesian posteriors fit well (R^2^=96.5%; Fig. 2 right) with low bias and imprecision (−0.15 and 1.07, respectively), and population PK model produced good fits of the observed cefepime concentrations (R^2^=87.0%, bias=0.53, imprecision=7.75; Fig. 2 left). The OFV change from the weight-adjusted, two-compartment model to final model was significant at −34 (p<0.05) (Table 1). Thus, a two-compartment model with weight and CrCL as covariate adjustment and allometric scaling was selected as the final PK model. The population parameter values from the final PK model are summarized in Table 2. Structural model and differential equations that define the population PK are listed in Supplemental Materials. The population parameter value covariance matrix can be found in Table 3. For the evaluation subset, Bayesian priors resulted in reasonably accurate and precise predictions (population R^2^=84.0%, Bayesian R^2^=90.2%; Fig. 3).

**Table 1.** Population Pharmacokinetic Model Builds and Comparisons.

| Models | -2LL | AIC | Change<br>in OFV | Population |  |  | Bayesian |  |  |
| --- | --- | --- | --- | --- | --- | --- | --- | --- | --- |
|  |  |  |  | Bias | Imprecision | R<sup>2</sup> | Bias | Imprecision | R<sup>2</sup> |
| **Base one** | 3223 | 3229 | - | 0.82 | 21.3 | 0.23 | -0.05 | 1.66 | 0.847 |
| **Base two** | 2994 | 3004 | 239 | 1.91 | 41.9 | 0.28 | 0.70 | 3.87 | 0.852 |
| **Two \*** | 2990 | 3000 | 4 | 2.17 | 68.4 | 0.61 | -0.18 | 1.12 | 0.965 |
| **Two \*\*** | 2966 | 2978 | 34 | 0.53 | 7.75 | 0.87 | -0.15 | 1.07 | 0.965 |

\*weight adjusted
\*\*weight adjusted and allometric scaler applied

**Table 2.**
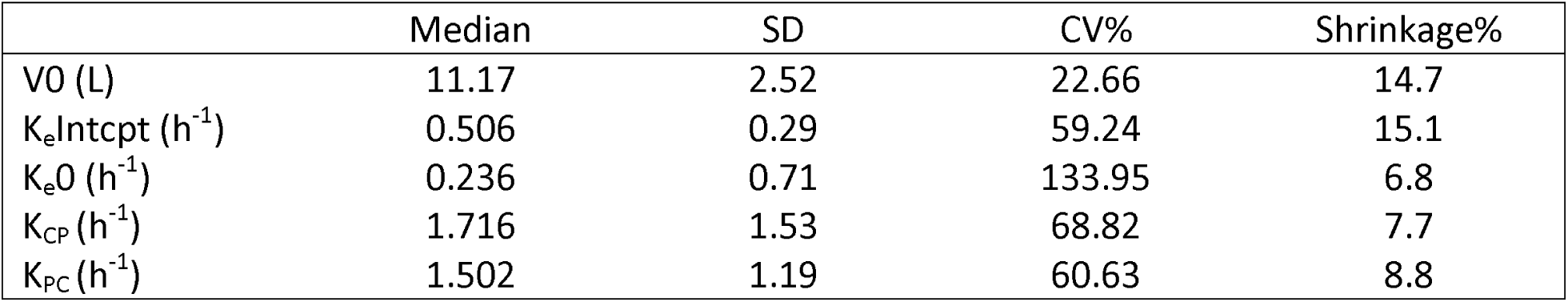
Population PK Parameter Estimates from the Final Model.

**Table 3.** Population Parameter Value Covariance Matrix for the Final Model.

|  | **V<sub>0</sub>** | **K<sub>e</sub>Intcpt** | **K<sub>e</sub>0** | **K<sub>CP</sub>** | **K<sub>PC</sub>** |
| --- | --- | --- | --- | --- | --- |
| **V<sub>0</sub>** | 6.366 | - | - | - | - |
| **K<sub>e</sub>Intcpt** | -0.106 | 0.085 | - | - | - |
| **K<sub>e</sub>0** | -0.238 | -0.152 | 0.499 | - | - |
| **K<sub>CP</sub>** | -1.576 | 0.005 | 0.145 | 2.354 | - |
| **K<sub>PC</sub>** | -0.680 | 0.027 | -0.188 | 1.441 | 1.414 |

**Figure 2.**
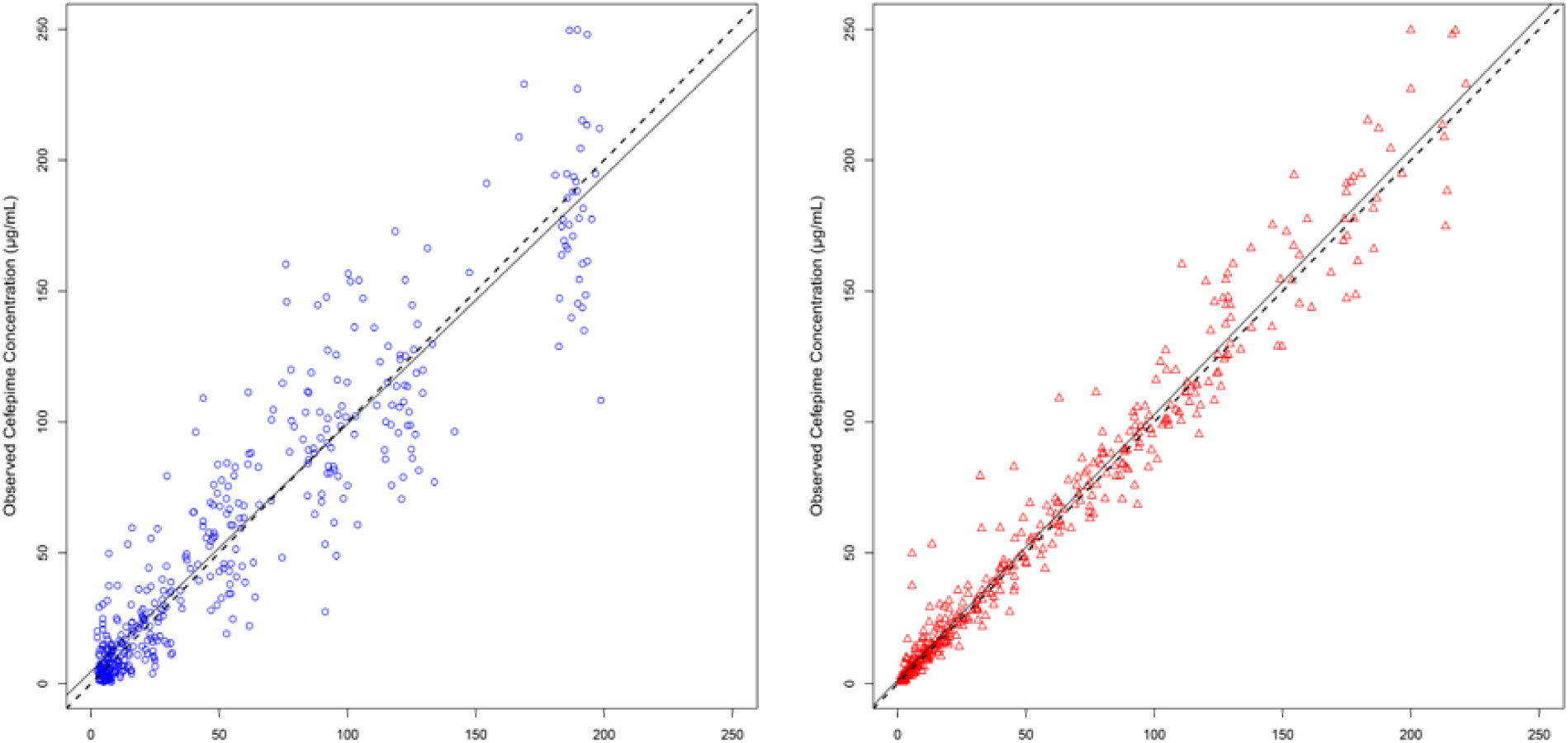
Goodness-of-fit Plots for Best-Fit Population Cefepime Model. Solid line denotes linear regression; dashed line denotes reference line For population predicted (blue), R^2^=0.87, slope=0.946 (95% CI, 0.912 to 0.981), bias=0.529, imprecision=7.75; for individual predicted (red), R^2^=0.965, slope=1.01 (95% CI, 0.996 to 1.03), bias=-0.148, imprecision=1.07

**Figure 3.**
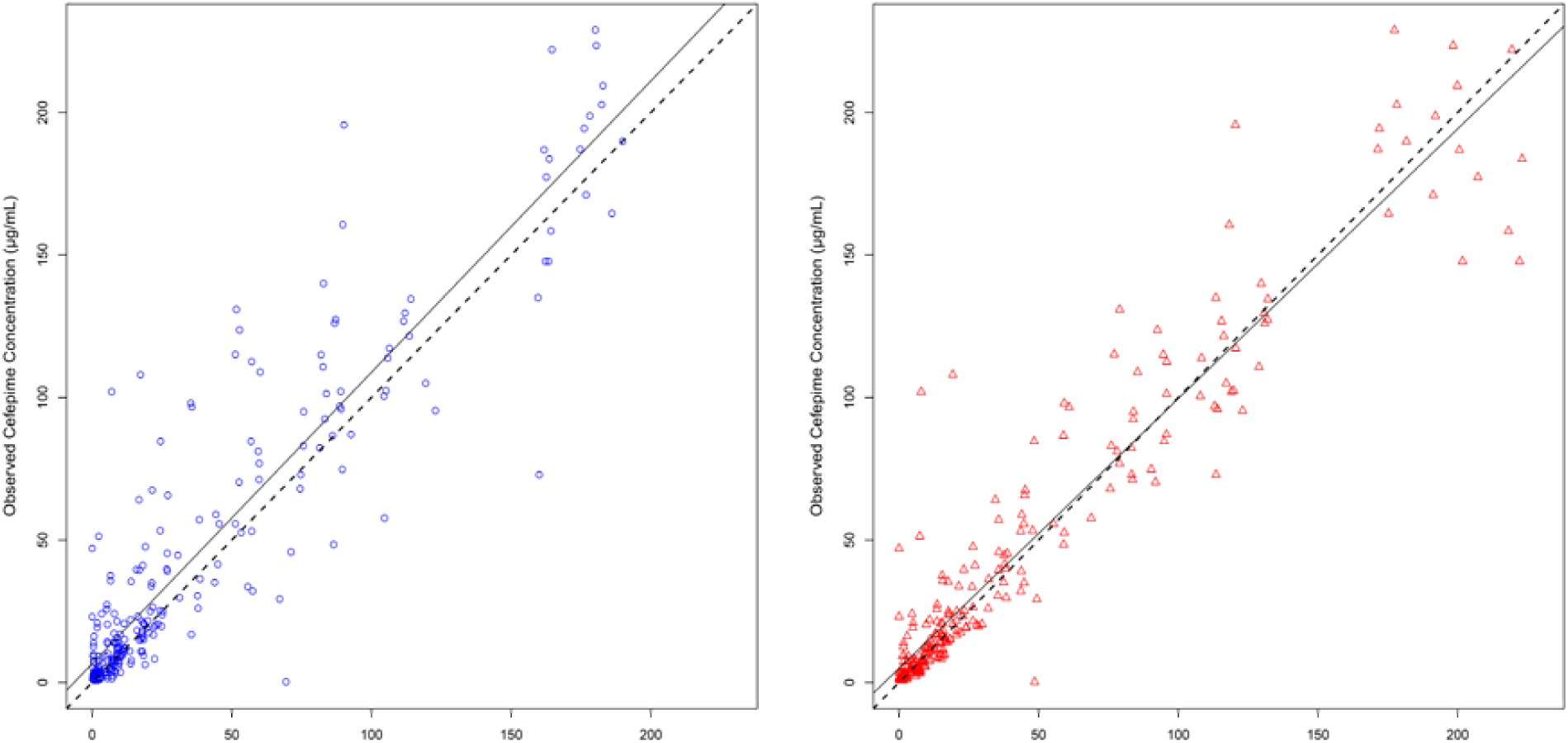
Goodness-of-fit Plots for Evaluation of Population Cefepime Model. Solid line denotes linear regression; dashed line denotes reference line For population predicted (blue), R^2^=0.84, slope=1.02 (95% CI, 0.967 to 1.08); for individual predicted (red), R^2^=0.902, slope=0.947 (95% CI, 0.908 to 0.985)

### 3.3 Simulation and PTA

Results of PTA analysis are shown in Table 4 and Figure 4 for the first 24 h of therapy. Two cefepime regimens were utilized to simulate PTA for adult and pediatric subjects. Cefepime dosage of 2 grams every 8 h infused over 30 minutes produced PTAs of >90% for MICs of 0.25-2 mg/L while a more than 2-fold drop of PTA was observed from MIC of 4 mg/L to 8 mg/L. The second cefepime regimen of 800 mg every 8 h infused over 30 minutes achieved PTA of >90% only at an MIC of 0.25 mg/L, PTA at 81.1% at MIC of 0.5 mg/L and performs poorly across subsequent higher doubling MICs.

**Table 4.** Probability of Target Attainment at Different Cefepime MICs (Maximum Recommended Dosages for Adults and Pediatrics) for the First 24 Hours of Therapy

| Cefepime regimen |  |  | Cefepime MICs (mg/L) |  |  |  |  |  |  |  |
| --- | --- | --- | --- | --- | --- | --- | --- | --- | --- | --- |
| Dose (mg) | Dosing interval (hr) | Infusion time (hr) | 0.25 | 0.5 | 1 | 2 | 4 | 8 | 16 | 32 |
| 2000 | 8 | 0.5 | 100% | 99.5% | 97.6% | 94.3% | 78.5% | 36.6% | 5.5% | 0.3% |
| 800 | 8 | 0.5 | 93.6 | 81.1% | 75.2% | 58.8% | 33.1% | 11.4% | 1.7% | 0.3% |

**Figure 4.**
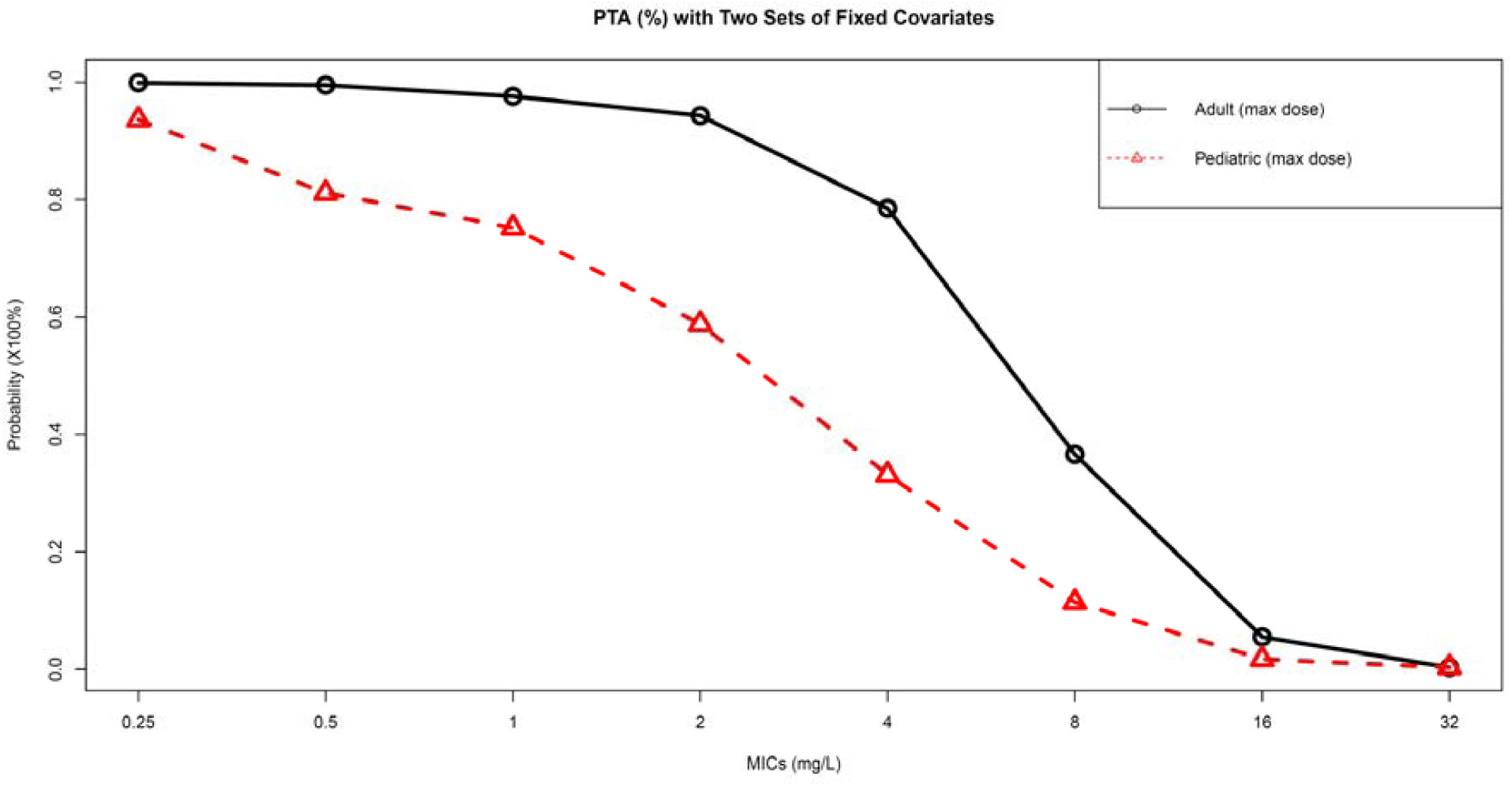
Probability of Target Attainment at Different Cefepime MICs.

## 4. Discussion

This study created a population and individual PK model for adult and pediatric patients and can serve as a Bayesian prior for precision dosing. When paired with a real time assay for cefepime, this model will allow for precise and accurate predictions of cefepime disposition via adaptive feedback control. In the absence of real time assays, these cefepime PK parameters facilitate more accurate population-based dosing strategies. Previous work by Rhodes et al. has shown an absolute difference of approximately 20% in survival probability across the continuum of achieving 0-100% *f*T_>MIC_ in adult patients with Gram-negative bloodstream infections, thus understanding the dose and re-dosing interval necessary to achieve optimal PK exposures should greatly improve clinical outcomes for patients treated with cefepime [5].

Individualized dosing and therapeutic drug monitoring (TDM) of β-lactam antibiotics (e.g. cefepime) are critically important to achieving optimal drug exposure (i.e. optimal *f*T_>MIC_ as the PK/PD target) and improving clinical outcomes [4, 21, 22]. Precision medicine has been named as a major focus for the National Heath Institute with $215 million invested [23], yet precision medicine has mostly focused on genomic differences [24, 25]. Precision dosing is an important facet of precision medicine, and renewed efforts in precision dosing in the real-world setting are being pursued [26]. Cefepime is a highly relevant example. While rigorous reviews and analyses are conducted during the development phase of an antibiotic, dose optimization is far less ideal for the types of patients who ultimately receive the drug. This is highlighted by the fact that although cefepime-associated neurotoxicity is rare. This serious and potentially life-threatening adverse event has been increasingly reported, yet few strategies exist for optimizing and delivering precision exposures [27, 28]. Lamoth et al. conducted a study to investigate the PK/PD threshold for cefepime-associated neurotoxicity [29]. Their final model predicted that a cefepime trough concentration of ≥22 mg/L (p=0.05) has a 50% probability of predicating neurotoxicity. In contrast, Rhodes et al. performed simulations from literature data and found that such threshold by Lamoth et al. is suboptimal as a predictive trough cutpoint [30]. Moreover, high intercorrelation amongst all PK parameters (i.e. AUC_SS_, C_MAX_, and C_MIN_) was observed by Rhodes et al., suggesting more work is needed to establish PK/TD profile for cefepime. Similarly, Huwyler et al. found a different trough (≥35 mg/L) as the predictive cefepime neurotoxic threshold, contrasting that of Lamoth et al. [31]. In addition to complications by these less-than-ideal PK/TD data, clinicians are left to treat patients with extreme age differences, organ dysfunction, and comorbid conditions affect antibiotic PK/PD [22]. These ‘real-world’ patients are often under-represented, and thus not well understood, from a PK/PD and PK/TD standpoint during the drug approval process. Bridging to the more typical patients that are clinically treated is important and central to the mission of Precision Medicine. Findings of this study can be used to guide cefepime dosing in these ‘real-world’ patients.

Several other studies have reviewed population cefepime PK. Sime et al. observed that patients with neutropenic fever had a mean clearance (CL) of 8.6 L/h, mean elimination half-life of 2.7 h [10]. Nicasio et al. studied 32 critically ill patients receiving intravenous cefepime for ventilator-associated pneumonia and observed that means of total CL and elimination half-life (as calculated from mean total CL and mean volume of distribution) were 7.6 L/h and 2.0 h, respectively [32]. The NCA conducted (in model development adult population) in our study produced similar results (CL, 7.59 L/h; elimination half-life, 2.98 h). Shoji et al. studied 91 pediatric patients and observed a mean of CL of approximately 1.86 L/h and a mean elimination half-life of 3.5 h (23). In our pediatric population, means of CL and elimination half-life were 3.1 L/h and 3.0 h, respectively. Our simulation findings are similar to those of Shoji et al. that a maximum pediatric cefepime dosing of 50 mg/kg every 8 h or 12 h did not adequately achieve optimal exposure to target higher MICs. Other studies also performed simulation for PTA with different cefepime regimens and renal functions, Tam et al. found that with a PD target of 67%, 2 grams every 8 h (30-minute infusion) achieved approximately 90% PTA for MIC of 8 mg/L in patients with CrCL of 120 mL/min while 2 grams every 12 h achieved barely above 80% PTA for MIC of 4 mg/L in the same population [33]. Nicasio et al. also conducted a simulation using a PD target of 50% in the critically ill with varying renal function. Maximum recommended dosage (2 grams every 8 h) in patients with CrCL between 50-120 mL/min achieved a PTA of 78.1% at MIC=16 mg/L; however, when the same regimen was infused over 0.5 h, the PTA achieved was significantly lower [32]. Collectively, these findings suggest that cefepime exposure is highly variable and may be clinically suboptimal in a large number of patients commonly treated with cefepime. These findings support the need of precision dosing and TDM for β-lactam antibiotics to reach optimal PK/PD target given the high variability in drug exposures.

Our study is not without limitations. Although a relatively large and diverse cohort was included in model development and evaluation, we did not specifically assess certain subgroups such as patients with morbidly obesity, severe renal dysfunction, etc. These conditions may require patient-specific models. Secondly, many studies to date included “real-world” patients with various disease sates (e.g. neutropenic fever, renal failure, sepsis, etc.); however, all studies were conducted under research protocol where doses, administration times, etc. were all carefully confirmed. Additional efforts will be needed to evaluate performance in clinical contexts.

## 5. Conclusions

A unified population model for cefepime in adults and pediatric populations was developed and demonstrated excellent performance on evaluation. Current cefepime dosages are often suboptimal, and population variability is high. Precision dosing approaches and real time assays are needed for cefepime to optimize drug exposure and improve clinical outcomes.

## Data Availability

Data are from primary manuscripts

## Acknowledgements

None

## Supplementary Materials

Differential equations for two-compartment population PK model with covariate adjustments are:

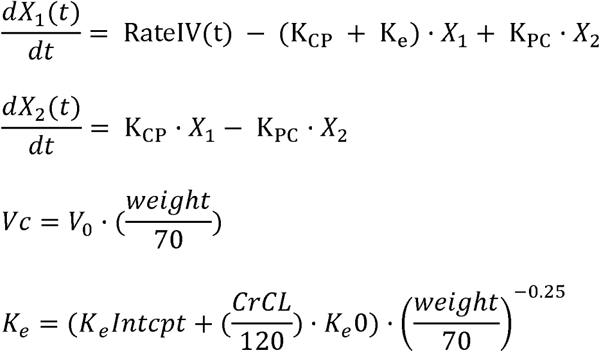

where Vc is the central cefepime volume of distribution, Ke is the elimination rate constant from the central compartment (h^-1^), KCP is the rate constant from central to peripheral compartment (h^-1^), and KPC is the rate constant from peripheral to central compartment (h^-1^).

